# Determinants of Hepatitis B Viral infection among pregnant women in Greater Monrovia, Liberia

**DOI:** 10.1101/2024.01.26.24301847

**Authors:** Henry Torbandu Kohar, George Asumah Adu, Henry Ofosu Addo, Edwin Afari, Ernest Kenu, Frederick Wurapa

## Abstract

**Background:** The global prevalence of hepatitis B virus infection is estimated to affect over 2 billion individuals, with a notable proportion ranging from 6% to 25% residing in the Sub-Saharan African region. The prevalence of hepatitis B virus (HBV) in Liberia, Guinea, and Sierra Leone has been estimated to be approximately 2%. However, current understanding of the actual risk factors associated with HBV in the Greater Monrovia remains unclear. Consequently, this study aimed to identify the factors associated with hepatitis B Viral infection among pregnant women residing in Greater Monrovia, Liberia.

**Methods:** An unmatched case control study of 141 cases of HBV infected pregnant women and 141 controls was conducted. Data on socio-demographic characteristics, lifestyle activities and medical related risk factors were collected for both groups with a structured questionnaire. Bivariate and multivariate analyses established associations between the HBV infection and risk factors studied.

**Results:** In all, 141 cases of HBV infected pregnant women and 141 controls participated in this study. The mean age of cases was 35.6 years (SD ±9.5) and 36.1 years (SD ±8.4) for controls. Low-income level <100 dollars [aOR 13.0 (4.48-37.82, p<0.001)], employment [aOR 0.04 (0.09-0.18) p<0.001], STI history [aOR 5.19 (1.68-16.02, p=0.004)] and living with a HBV infected person [aOR 35.11 (4.24-58.90), p=0.001] were factors associated with HBV infection among pregnant women in Greater Monrovia.

**Conclusion:** The risk factors of HBV infection among pregnant women in Greater Monrovia were engagement in formal employment, low-income level, history of sexually transmitted infections (STIs), and residing with an individual infected with hepatitis B virus (HBV). Control efforts by designated state institutions should include advocacy and awareness creation on HBV status identification, safe sex, and vaccination of uninfected individuals.

## Introduction

Hepatitis B (HBV) is a viral infection that targets the liver and has the ability of causing both acute and chronic illnesses. According to the World Health Organization in 2019, hepatitis B infection accounted for around 820,000 fatalities, primarily attributed to cirrhosis and hepatocellular carcinoma, which is a kind of primary liver cancer (1,2). The transmission of the virus typically occurs through vertical transmission from mother to child (MTCT) during the process of childbirth, during early infancy, and through contact with blood or other bodily fluids during sexual intercourse with an infected partner, injections, or exposure to contaminated sharp tools (3,4). Hepatitis B viral infection is of public health concern (1,5).The prevalence of chronic infections was estimated to be at 296 million individuals in 2019, with an annual incidence of 1.5 million new infections (6,7). Amongst these cases, it was observed that 68% resided in Africa and Western Pacific Asia (8,9). Moreover, it has been reported that a significant proportion, ranging from 5% to 15% of individuals residing in underdeveloped nations and countries with low income are persistently infected with the hepatitis B virus (10,11). In addition, the World Health Organization (WHO) reveals that hepatitis accounted for 1.34 million fatalities in 2015, a figure comparable to tuberculosis-related deaths but exceeding those attributed to HIV (12). Furthermore, a substantial number of children under the age of five, specifically 1.8 million, are currently afflicted with hepatitis B virus infection due to the high prices of HBV vaccine (13,14).

Hepatitis B Virus disease is more prevalent in Sub-Sahara Africa; about 6 to 25 percent Sub Saharan Africans are infected with the disease (15,16). Greater Monrovia region and Liberia has a prevalence of about 2% (17). HBV among pregnant women poses a lot of threats to mother and fetus, leading to complications such as preterm delivery, low birth weight, threatened post-partum hemorrhage and other associated complications. It is estimated that up to 40% of Hepatitis B virus prevalence can be attributed to mother to child transmission, and the core factor for chronic HBV infection (18,19). Several studies have associated HBV infections to history of multiple sexual partners, previous blood transfusions, caring or living with HBV positive family member, tattooing, illicit drug use, accidental pricking with sharps and having unsafe abortions among others (20,21). Presently, knowledge about the key risk factors is limited in Greater Monrovia and Liberia, therefore, this disease has not gained full national commitment and support from the Ministry of Health and other developing partners in Liberia. Greater Monrovia, being a business region, has a lot of its inhabitants constantly engaging in many risky behaviors which could expose them to the disease. Hence, this study sought to explore factors associated with HBV infection among pregnant women in Greater Monrovia to improve knowledge for enhanced control efforts.

## Methods

### Study Design

This study adopted an unmatched case-control study using quantitative research methods from August 2016 to July 2017. Data on Hepatitis B serum Agglutination (HbsAg) test results of both cases and controls as well as data on socio-demographic, previous medical exposures and lifestyle factors were collected from participants in diverse locations as deemed appropriate and convenient by participants.

### Study area

This study was conducted in Greater Monrovia, the capital and commercial city of Liberia Greater Monrovia comprises all the ethnic groups in Liberia and has a population of 1.5 million (Figure 1). It is bordered to the south by the Atlantic Ocean, the North by Paynesville, to the east by the Duazon community and to the west by Bushrod Island (Figure 1).

**Figure 1:**
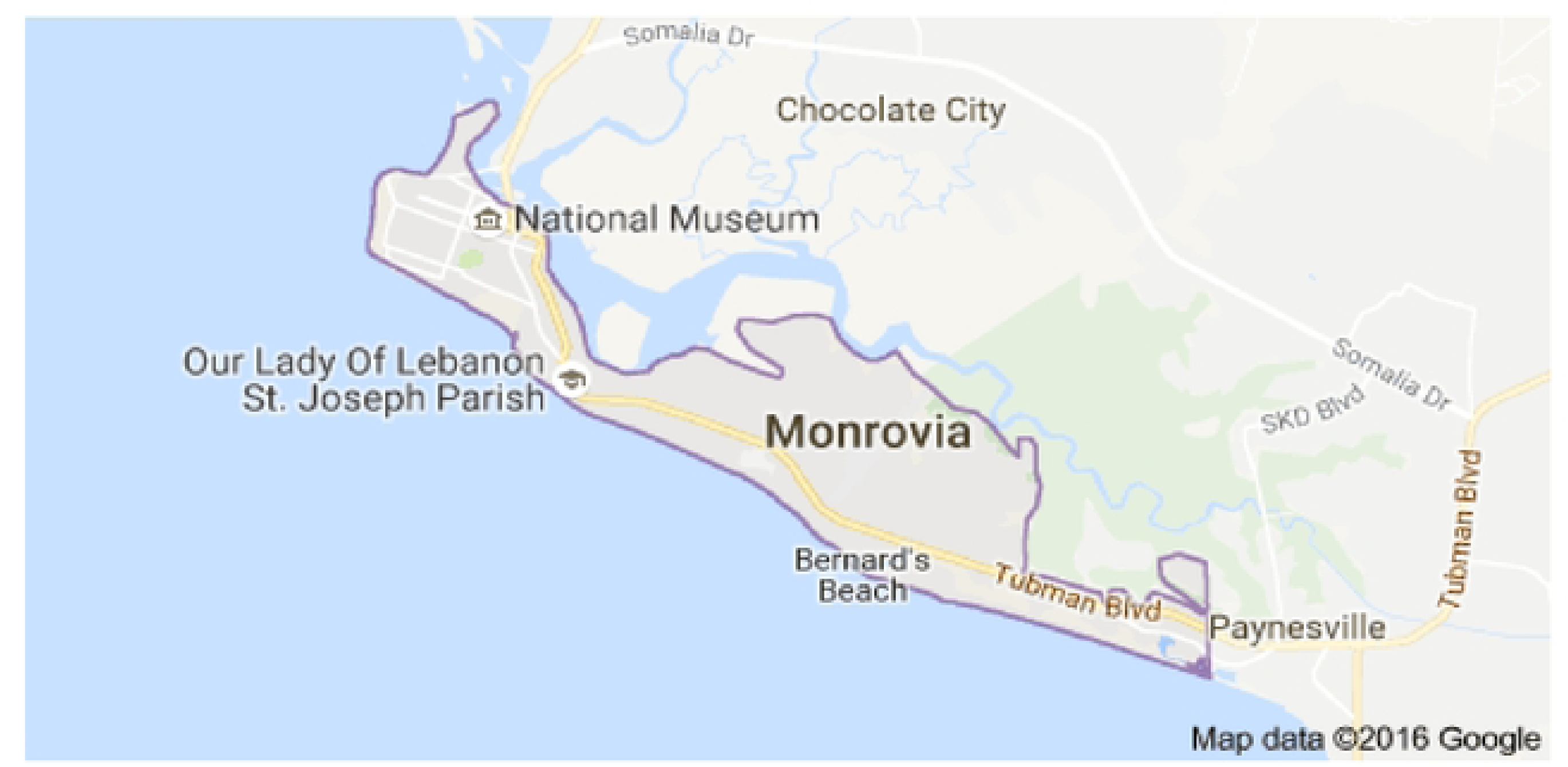
Map of study area

### Study population

This study involved pregnant women between the ages of 18-50 years registered at an Antenatal Clinic (ANC) in Greater Monrovia between the period of January 2015 to March 2017.

### Case Definitions Case

A case was defined as an ANC registrant between the ages of 18-50 years, who resided in Greater Monrovia and attended ANC from January 2015–March 2017 and tested positive to HbsAg test.

### Control

A control is defined as an ANC registrant between the ages of 18-50 years, who resided in Greater Monrovia and attended ANC from January 2015–March 2017 and tested negative to HbsAg test.

### Sample size determination

To estimate the sample size for this study, a stat calculator in Epi info version 7 was used. It was assumed that 20% of the study population are exposed to the risk factors of HBV infection (22). A detection of an odds ratio of 2.5 with an 80% power is to be achieved with a 95% confidence level (5% significance). A ratio of 1:1 case and control was used and since the prevalence of several of the known risk factors for HBV, is unavailable for Liberia, the figure used as the least exposure prevalence was multiple sexual contacts which was derived from by a study conducted by (23). A total of 141 cases and 141 controls was derived and used for this study.

### Sampling procedure

A multi-staged sampling was applied to select participants for this study. First of all, health facilities that provide ANC services were grouped into three based on facility ownership as follows: private, public and faith-based facilities. Table of random numbers was employed to select one facility from each group of facilities. The sampled facilities were John F. Kennedy Medical Centre for private hospitals, Eternal Love Winning Africa (ELWA) and James N. Davis (JFD) for the Faith-based and Government hospitals respectively. Based on proportional to size sampling, 40% of the sample size (56 cases and corresponding controls) were drawn from the John F. Kennedy Memorial Hospital accounted for approximately, ELWA also accounted for 30% (43 cases and corresponding controls), and a government hospital, and James N. Davis (JFD) contributed approximately 30% (42 cases and corresponding controls). A line list of pregnant women who tested positive for the HbsAg test was then generated from the ANC registration records for a period spanning January 2015 to March 2017. Finally, a table of random numbers was used to select respondents for the study. Same procedure was applied to the selection of controls in this study.

### Data Collection

A semi-structured questionnaire consisting of questions on demographic characteristics, lifestyle and some previous medical exposures was administered face to face to the participants in their preferred places or via telephone in few instances where physical interviews was not possible. The questionnaire was pretested at Redemption Hospital in Greater Monrovia for reliability and efficiency before actual data was collected. The questionnaire was administered in English language as it is the official main language of communication of people in Monrovia. Help was also sought from an interpreter where participant speaks different tongue.

### Data analysis

The raw data was inputted into Microsoft Excel spreadsheet (version 2010) and analyzed using Stata 13.0 software. Data was summarized as frequencies, proportions, means and medians where appropriate. Logistic regression analysis at p-value (<0.05) significance level was used to assess association between the risk factors associated with HBV infection.

### Ethical consideration

This study received ethical approval from the University of Liberia – Pacific Institute for Research & Evaluation Institutional Review Board (UL-PIRE IRB Assurance number FWA00004982). Formal permission was sought from the heads of John F. Kennedy Memorial Hospital, Eternal Love Winning Africa (ELWA) and James N. Davis Memorial Hospital. Written consent was obtained from all study participants before interviews were carried out. Study participants were made to understand their participation in the study was voluntary and could opt out of the study should they feel uncomfortable at any point in the study.

## Results

### Socio- demographic characteristics of pregnant women, Monrovia in Liberia, 2017

The mean age of cases was 35.6 years (SD ±9.5) and 36.1 years (SD ±8.4) for controls. More than a third of cases 39.0% (55/141) and controls 34.8% (49/141) had no formal education. Concerning employment, 5.0% (7/141) of cases compared with 21.3% (30/141) of controls were employed in the formal sector (Table 1).

**Table 1:** Socio-demographic characteristics of participants.

| Variables | Cases | Frequency (%) | Control | Frequency (%) |
| --- | --- | --- | --- | --- |
| Age (Mean) | 35.6 (SD $\pm 9.5$) | | 36.1 (SD $\pm 8.4$) | |
| **Educational status** |  |  |  |  |
| No Formal education | 55 | 39.0 | 49 | 34.8 |
| Primary/JHS | 26 | 18.4 | 27 | 19.1 |
| Secondary | 45 | 32.0 | 51 | 36.2 |
| Tertiary | 15 | 10.6 | 14 | 9.9 |
| **Employment** |  |  |  |  |
| Formal Sector | 7 | 5.0 | 30 | 21.3 |
| Informal Sector | 127 | 90.0 | 100 | 70.9 |
| Unemployed | 7 | 5.0 | 11 | 8.0 |
| **Marital Status** |  |  |  |  |
| Married | 113 | 80.1 | 101 | 71.6 |
| Single | 28 | 19.9 | 40 | 28.4 |

### Socio-demographic characteristics and medical risk factors associated with hepatitis B viral infection

At crude level, earning an income of <100USD [cOR 6.32 (3.29-12.54), P<0.001], having undergone major surgical procedure [cOR=4.71 (2.16-11.07) P<0.00)], ever been diagnosed of an STI [cOR=4.7 (2.16-11.07) P=0.020] and having a history of abortion [cOR= 3.44 (1.42-9.18), P=0.003] increased the odds of HBV infection. On the other hand, been engaged in a formal sector employment [cOR= 0.19 (0.08-0.43), P<0.001] reduced the odds of HBV infection in this study (Table 2).

**Table 2:** Bivariate analysis of socio-demographic and medical characteristics of pregnant women in Greater Monori Liberia.

|  | **Hepatitis B Disease** |  | **cOR (95% CI)** | **P value** |
| --- | --- | --- | --- | --- |
|  | **Cases** | **Controls** |  |  |
| **Age of PW** |  |  |  | 0.10 |
| ≥ 30 years | 89 (63.1) | 102 (72.3) | 0.66 (0.38-1.12) |  |
| < 30 years | 52 (36.9) | 39 (27.7) | 1.0 |  |
| **Marital Status of PW** |  |  |  | 0.10 |
| Single | 28(19.9) | 40 (28.4) | 0.63 (0.35-1.13) |  |
| Married | 113 (80.1) | 101 (71.6) | 1.0 |  |
| **Education of PW** |  |  |  | 0.55 |
| ≥SHS | 60 (42.6) | 65 (46.1) | 0.87 (0.52-1.42) |  |
| <SHS | 81(57.4) | 76 (53.9) | 1.0 |  |
| **Employment status of PW** |  |  |  | <0.000 |
| Formal Sector | 7 (6.0) | 30 (23.0) | **0.19 (0.08-0.43)** |  |
| Informal Sector | 127 (94.0) | 100 (77.0) | **1.0** |  |
| **Income level of PW** |  |  |  | <0.000 |
| <100USD | 62(44.9) | 16 (11.4) | **6.32 (3.29-12.54)** |  |
| ≥100USD | 76 (55.1) | 124 (88.6) | **1.0** |  |
| **Blood History of Blood transfusion** |  |  |  |  |
| Yes | 70 (50.4) | 64 (45.4) | 1.22 (0.74-1.0) | 0.41 |
| No | 69 (49.6) | 77 (54.6) | 1.0 |  |
| **Major surgical procedure** |  |  |  |  |
| Yes | 37(26.4) | 10 (7.1) | **4.71 (2.16-11.07)** | <0.000 |
| No | 103 (73.6) | 131 (92.9) | **1.0** |  |
| **Ever diagnosed an STI** |  |  |  |  |
| **no data for this** |  |  |  |  |
| Yes | 72(53.2) | 52(37.0) | **1.79 (1.08-2.98)** | **0.020** |
| No | 67(46.8) | 87(63.0) | **1.0** |  |
| **Ever done Abortion** |  |  |  |  |
| Yes | 24 (17.3) | 8 (5.7) | **3.44 (1.42-9.18)** |  |
| No | 115 (82.7) | 132 (94.3) | **1.0** | **0.003** |

### Lifestyle risk factors associated with HBV infection

Participants who had body tattoo and/or piercing [cOR= 2.46 (1.45-4.19), P=0.004], having multiple sexual partners [cOR=17.06 (2.52-75.0), p<0.001], having ever lived with HBV infected person [cOR 17.31 (5.48-67.79), P<0.001] and use of illicit drugs increased the odds of HBV infection (Table 3).

**Table 3:**
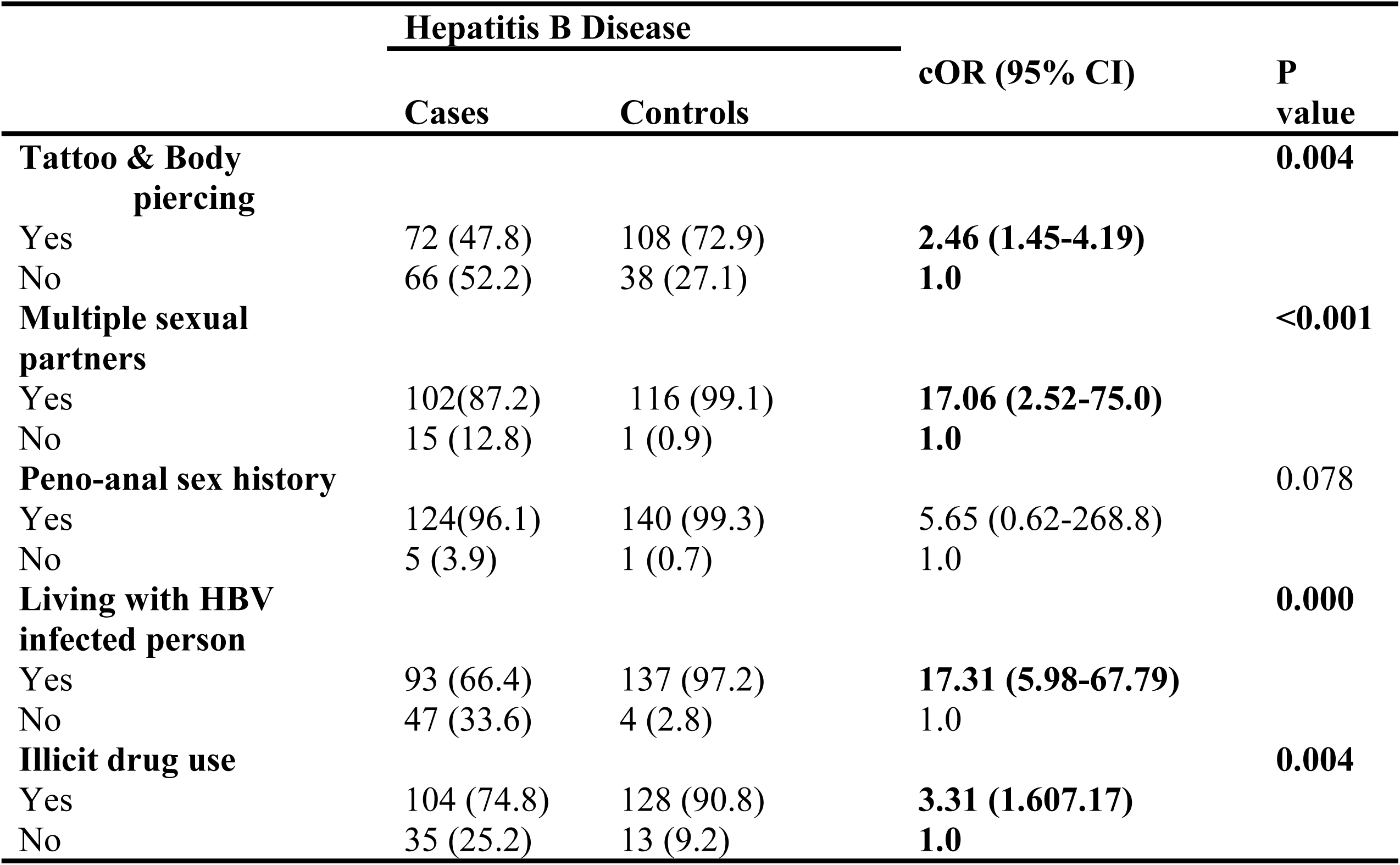
Lifestyle characteristics of pregnant women and its association with HBV infection.

### Factors associated with HBV among pregnant women, Greater Moronvia, 2017

After adjusting for confounders, employment status, income level, history of STI and living with an HBV infected person remained significantly associated with HBV infection. Pregnant women who earned less than USD 100 showed higher odds [aOR=13.0 (4.48-37.82, p< 0.001) of HBV infection than those who earned ≥USD 100. Pregnant women who were employed in the formal sector had a lower odd [aOR=0.04 (0.09-0.179), p-value <0.001] of HBV infections than those who were employed in the informal sector. Regarding STIs, pregnant women who reported history of STI had higher odds [aOR=5.19 (1.68-16.02), p= 0.004] of been diagnosed HBV positive than those who had no STI history. Also, having a history of living with an HBV infected person rendered pregnant women more prone [aOR=35.11 (4.24-58.9), p=0.001] to HBV infection than those with no history (Table 4).

**Table 4:** Multivariate analysis of determinants and its association with HBV among pregnant women.

| Variable | HBV diseases |  | Adjusted Odd ratio | P value |
| --- | --- | --- | --- | --- |
|  | Cases | Controls | (95%CI) |  |
| Income level of PW |  |  |  |  |
| <100USD | 76 (55.07) | 124 (85.71) | 13.0 (4.48-37.82) | <0.001 |
| ≥100USD | 62(44.93) | 16 (14.29) | 1.0 |  |
| Employment status of PW |  |  |  |  |
| Unemployed | 132 (93.61) | 104 (73.76) | 0.04 (0.09-0.179) | <0.001 |
| Employed | 9 (6.39) | 37 (26.24) | 1.0 |  |
| Major surgical procedure |  |  |  |  |
| Yes | 103 (73.57) | 131 (92.91) | 4.06 (0.731-22.59) | 0.109 |
| No | 37(26.43) | 10 (7.09) | 1.0 |  |
| Illicit drug use |  |  |  |  |
| Yes | 104 (74.83) | 128 (90.78) | 4.02 (0.39-40.44) | 0.237 |
| No | 35 (25.17) | 13 (9.22) | 1.0 |  |
| **Tattoo & Body piercing** |  |  |  | 0.535 |
| Yes | 72 (47.82) | 108 (73.98) | 0.86 (0.24-2.14) |  |
| No | 66 (52.18) | 38 (26.02) | 1.0 |  |
| **Multiple sexual partners** |  |  |  | 0.425 |
| Yes | 102(87.18) | 116 (99.15) | 2.81 (0.22-35.55) |  |
| No | 15 (12.82) | 1 (0.85) | 1.0 |  |
| **Living with HBV infected person** |  |  |  | <0.001 |
| Yes | 93 (66.42) | 137 (97.17) | **35.11 (4.24-58.9)** |  |
| No | 47 (33.58) | 4 (2.83) | 1.0 |  |
| **Ever diagnosed an STI** |  |  |  | **0.004** |
| Yes | 75(53.2) | 52(37.0) | **5.19 (1.68-16.02)** |  |
| No | 66(46.8) | 87(63.0) | 1.0 |  |

## Discussion

Hepatitis B virus infection is one of the world’s leading public health challenges, causing significant morbidity and mortality (24). A newborn infant whose mother is positive for both HBsAg and HBeAg has a 90% chance of developing chronic infection by the age of 6 (24). Hepatitis B virus infection among pregnant women pose a serious public threat to both mother and baby and must be given the needed attention. It is on this premise that this study sought to determine factors associated with Hepatitis B Virus infection among pregnant women of Greater Monrovia.

This study found an association between Hepatitis B virus infection and factors including informal sector employment, low-income status, having a history of Sexually Transmitted Infections (STIs), and living with a Hepatitis B Virus infected person.

### Socio-demographic factors and medical history

Current study indicates reduced odds of contracting HBV among pregnant women who were in the formal sector employment than those who were in the informal sector employment. This can be attributed to their improved economic and educational status leading to increased knowledge and awareness of HBV as well as having access to good medical care. Present findings agrees to a study conducted in Ethiopia as pregnant women who were unemployed were 8 times at risk of HBV infection than those who were employed (25). A recent study on the knowledge of hepatitis B virus among pregnant women in the Ningo-Prampram District in Ghana found a strong association between educational level and positivity to hepatitis b virus (26). This study also found an association between income status and HBV, as participants who earn <100USD had higher odds of developing HBV infection than those who earn ≥100USD. This agrees with a study conducted in Brazil which indicates a higher prevalence of HPV infection among individuals of lower socioeconomic status (27). Another study in Japan also confirms association between socioeconomic status and hepatitis B virus prevalence (28). Additionally, pregnant women who reported ever having sexually transmitted infection were more likely to be HBV infected than those who had never had any STIs.

### Lifestyle factors associated with HBV infection

The investigation delved into lifestyle-related factors, elucidating their association with HBV infection. Living with a Hepatitis B infected person was significantly associated with HBV infection among pregnant women in in this study. The virus can spread through contact with infected body fluids such as blood, saliva, vaginal fluids, and semen. Living with an infected person exposes you to these fluids thereby increasing risk of transmission. This concurs with many recent findings (24,26). A study conducted by Freitas et al. (2014) in Central Brazil also observed that pregnant women who resided with or had previous exposure to an individual infected with Hepatitis B had a higher likelihood of acquiring HBV infection compared to those who had no history of living with a Hepatitis B-infected individual. It is evident that pregnant women who had previously resided with an individual infected with Hepatitis B exhibited increased likelihood of being diagnosed as HBV positive, in comparison to those who had never cohabitated with a Hepatitis B infected individual (30,31). Another study conducted in Northwestern Ethiopia Molla et al. (2015) validates the findings of current study as it also observed that pregnant women who had previously resided or were now residing with an individual infected with Hepatitis B exhibited a greater likelihood of experiencing adverse outcomes related to HBV compared to pregnant women who had never cohabited with a Hepatitis B infected individual. Another study in Tanzania also reported similar findings (33).

## Conclusion

This study provides valuable insights into the complex interrelationships between socio-demographic, medical, and lifestyle factors influencing HBV infection among pregnant women in Greater Monrovia, Liberia. The study found that several risk factors significantly associated with Hepatitis B infection among pregnant women who registered for antenatal care in Greater Monrovia. These factors included having formal employment, having a low-income level, having a history of sexually transmitted infections, and living with a person infected with Hepatitis B virus. The multifactorial nature of HBV transmission underscores the importance of tailored interventions focusing on education, healthcare accessibility, behavioural modifications, and targeted screening programs to mitigate the burden of HBV among this vulnerable population. The Ministry of Health of Liberia and developing partners must enhance interventions aimed at preventing and controlling sexually transmitted infections (STIs), including initiatives focused on creating awareness on STI prevention and control as well as promoting HBV vaccination among uninfected individuals.

## Data Availability

All relevant data are within the manuscript and its Supporting Information files

## Acknowledgment

The authors would like to thank all the health facilities and participants who availed themselves for this study.

